# Right Ventricular Work and Pulmonary Capillary Wedge Pressure in Heart Failure with Preserved Ejection Fraction

**DOI:** 10.1101/2024.09.16.24313782

**Authors:** Kuan-Chih Huang, Ting-Tse Lin, Cho-kai Wu, Lung-Chun Lin, Lian-Yu Lin

## Abstract

**Background:** Symptoms of heart failure with preserved ejection fraction (HFpEF) are closely related to elevated pulmonary capillary wedge pressure (PCWP) during exercise. Understanding right ventricular (RV) myocardial work, using RV pressure–strain loops to assess RV function in HFpEF, is lacking. The study aims to evaluate the effectiveness of right ventricular myocardial work parameters in diagnosing HFpEF and their correlation with pulmonary capillary wedge pressure during exercise.

**Methods:** The study included patients who underwent invasive cardiopulmonary exercise tests, measuring pressures at rest and during exercise to identify HFpEF. Echocardiography assessed left and right ventricular parameters. RV myocardial work was calculated using strain-rate and pressure curves, matched with ECG data. RV global constructive work (RV GCW), RV global work index (RV GWI), RV global wasted work (RV GWW), and RV global work efficiency (RV GWE) were analyzed and compared with invasively measured PCWP at rest and peak exercise.

**Results:** Forty-one patients with adequate data were enrolled, with 21 diagnosed with HFpEF. No significant differences in various echocardiographic parameters were found between HFpEF and non-HFpEF groups, except higher post-exercise PCWP and mean pulmonary artery pressure in HFpEF patients. HFpEF patients had higher RV GWW and lower RV GWE. RV GWW and RV GWE had higher predictive ability for HFpEF diagnosis compared to other echocardiographic parameters. RV GCW (r = 0.504, P = 0.001) and RV GWW (r = 0.621, P < 0.001) correlated with post-exercise ΔPCWP and exercise PCWP, with RV GWW independently associated with both after adjustment for confounding factors.

**Conclusions:** RV GWW is a novel predictive parameter that provides a better explanation of RV performance regarding post-exercise ΔPCWP than other standard echocardiographic parameters in HFpEF.

**WHAT IS NEW?:**

1. Right ventricular global wasted work (RV GWW) is identified as a novel predictive parameter in heart failure with preserved ejection fraction (HFpEF), providing a more accurate explanation of right ventricular (RV) performance regarding post-exercise changes in pulmonary capillary wedge pressure (ΔPCWP) compared to standard echocardiographic parameters.
2. study highlights the significant correlation of RV GWW with post-exercise ΔPCWP and exercise PCWP, demonstrating its independent association after adjusting for confounding factors.
3. myocardial work, assessed using RV pressure–strain loops from invasive cardiopulmonary exercise tests and echocardiography, offers valuable insights into the role of RV in exercise intolerance and dyspnea among HFpEF patients.

**WHAT ARE THE CLINICAL IMPLICATIONS?:**

1. RV GWW serves as a superior predictive marker for diagnosing HFpEF, independently correlating with post-exercise ΔPCWP. This enhances the understanding of RV contribution to exercise intolerance and dyspnea in HFpEF, potentially aiding in more accurate diagnosis and tailored management strategies.
2. RV GWW analysis in clinical practice may improve the diagnostic precision and management of HFpEF, optimizing treatment strategies for patients with unexplained dyspnea and suspected HFpEF.

## INTRODUCTION

Heart Failure with Preserved Ejection Fraction (HFpEF) has surpassed Heart Failure with Reduced Ejection Fraction (HFrEF) in terms of prevalence. However, owing to its heterogeneous etiology, the diagnosis of HFpEF is more challenging ^1^. Cardiologists rely on clinical demographics, laboratory biomarkers, comprehensive echocardiography and functional studies for disease stratification. Although pulmonary hypertension and right ventricular dysfunction are anticipated in HFpEF ^2–4^, invasive Cardiopulmonary Exercise Test (iCPET) is still frequently needed to unmask the elevated left ventricular (LV) filling pressure in equivocal patients ^5,6^. Patients presented as dyspnea of unknown etiology could have normal pulmonary capillary wedge pressure (PCWP) at rest but suffer from increases in PCWP during exercise. The difference between PCWP measured during exercise and rest (ΔPCWP) is an important indicator of heart function, and a higher ΔPCWP was associated with a higher mortality rate ^7,8^. Our previous work revealed that exercise left atrial conduit strain was highly associated with ΔPCWP ^9^; however, rest echocardiography parameters predicting such hemodynamic change is lacking.

Myocardial work is a recent emerging concept that integrates information on pressure and myocardial strain throughout the entire cardiac cycle ^10,11^. In addition to the overall ventricular work index, this concept further delineates the notions of constructive work, wasted work, and work efficiency. Unlike global longitudinal strain and ejection fraction, which are limited by afterload dependency, myocardial work is considered a promising tool to detect incipient LV systolic dysfunction in aortic stenosis ^12^. Due to the thinner walls and lower ventricular elastance of the right ventricle (RV), RV longitudinal strain is even more afterload dependent ^13,14^. Recently, a proof-of-concept study demonstrated a significant correlation of RV myocardial work (RVMW) with invasively measured stroke volume and stroke volume index in a HFrEF population ^15^. Such technique should have considerable potential for detecting early RV dysfunction in patients with HFpEF.

Since the currently available non-invasive method for assessing myocardial work is still vendor-specific ^16^, and because right ventricular pressures are naturally lower than left ventricular pressures, the margin of error when estimating RV pressures using modified Bernoulli equation could impacts the results more in HFpEF than in HFrEF or pulmonary hypertension. The aim of this study is to design a semi-invasive RVMW method by combining the pressure recordings from Swan-Ganz catheter with strain results via echocardiography. We also investigate relationship between the values of RVMW and the results of iCPET for better understanding of the RV pathophysiology in HFpEF.

## MATERIAL AND METHODS

### Study population

Patients presenting with dyspnea of unknown etiology were recruited for this study upon the diagnosis of HFpEF. HFpEF was defined according to the 2016 ASE/EACVI HF criteria, characterized by the following criteria: (1) typical symptoms and signs of heart failure; (2) left ventricular ejection fraction (LVEF) ≥50%; (3) elevated NT-proBNP levels (≥125 pg/mL); and (4) echocardiographic evidence of structural abnormalities (left atrial volume index >34 mL/m² or left ventricular mass index ≥115 g/m² for men and ≥95 g/m² for women) or functional abnormalities (E/e’ ratio ≥13 and mean e’ velocity of septal and lateral wall <9 cm/s). Exclusion criteria included chronic renal failure (creatinine >250 μmol/L), significant hepatic disease, significant coronary artery disease (≥70% coronary artery stenosis without intervention or positive stress test), secondary hypertension, pericardial disease, significant valvular heart disease (>mild stenosis and >moderate regurgitation), cancer, cor pulmonale, congenital heart disease, left-to-right shunt, myocardial infarction within 60 days, high-output heart failure, and chronic atrial fibrillation. This study complied with the Declaration of Helsinki and received approval from the institutional review board of the National Taiwan University Hospital (201908057RINC). Informed consent was obtained from all participants prior to inclusion in the study. The patient enrollment algorithm is shown in Supplemental Figure 1. For subjects (N=41) with complete echocardiography and iCPET examination, HFpEF (N=21) diagnosis was confirmed if resting pulmonary capillary wedge pressure (PCWP) was ≥15 mm Hg or exercise-induced PCWP was ≥25 mm Hg.

### Invasive cardiopulmonary exercise test

A 7-F Swan–Ganz catheter (Biosensors International) was inserted into the pulmonary artery via the internal jugular vein sheath. Before and after a constant 20-W workload for 6 min on an electromagnetically braking cycle ergometer (Ergometrics ER800, Ergoline GmbH), patients underwent right cardiac catheterization at rest and during supine exercise ^17,18^. Transducers were zeroed at the mid-axilla. Right atrial (RA) pressure (RAP), pulmonary artery (PA) pressure (PAP), and PCWP were recorded at end-expiration. Mean RAP and PCWP were recorded at mid A wave. Patients met the criteria of resting PCWP >15mmHg and post-exercise PCWP >25mmHg were considered as HFpEF in the present study.

### Echocardiography

The 2D echocardiographic data set was acquired right before the invasive cardiopulmonary exercise test using a CX50 xMATRIX system equipped with an S5-1 transducer (Philips Medical Systems, Andover, MA) and stored digitally for offline analysis with dedicated software (TTA 2.3 Cardiac performance analysis, TomTec Imaging Systems, Unterschleißheim, Germany). LV apical 4-chamber, 2-chamber, and 3-chamber views were used to calculate the left ventricular end-diastolic volume (LVEDV), left ventricular end-systolic volume (LVESV) as well as the consequent left ventricular stroke volume (LVSV), left ventricular ejection fraction (LVEF), left ventricular global longitudinal strain (LVGLS), left atrial volume (LAV), left atrial reservoir strain (LASr), left atrial conduit strain (LASc) and left atrial booster strain (LASb). RV focused views were analyzed for RV end-diastolic area (RVEDA), RV end-systolic area (RVESA), RV fractional area change (RVFAC), RV global longitudinal strain (RVGLS), RV free wall strain (RVFWS) and tricuspid annular plane systolic excursion (TAPSE). Doppler and tissue doppler echocardiograms were used to calculate early (E) and late (A) diastolic transmitral velocities and mean E/e’ ratios. All the echocardiographic analyses were performed by an independent cardiologist who did not involve the image acquisition and was blind to the clinical data.

### Calculation of right ventricular myocardial work

The RVGLS derived strain rate-to-time curves were exported as .xml files via the dedicated software. A Swan-Ganz catheter was delivered into the RV apex during ICPEPT and the RV pressure curves were exported as .xml files along with electro-cardiography (ECG) tracings using a commercially available hemodynamic recording system (Mac-Lab Cardiac Cath Lab Physiological Recording System, GE Healthcare, Milwaukee, U.S.A.). Because the ventricular pressure was recorded but not estimated, we did not use the doppler defined timing (aortic valve opening, aortic valve closure, mitral valve opening, mitral valve closure) to match the RV strain-rate and RV pressure curves; instead, we used the ECG to conjugate the RV strain-rate and RV pressure curves (Figure 1). RV Global work index (GWI), global constructive work (GCW), global waste work (GWW) and global work efficiency (GWE) were defined as their left ventricular counterparts ^19^.

**Figure 1.**
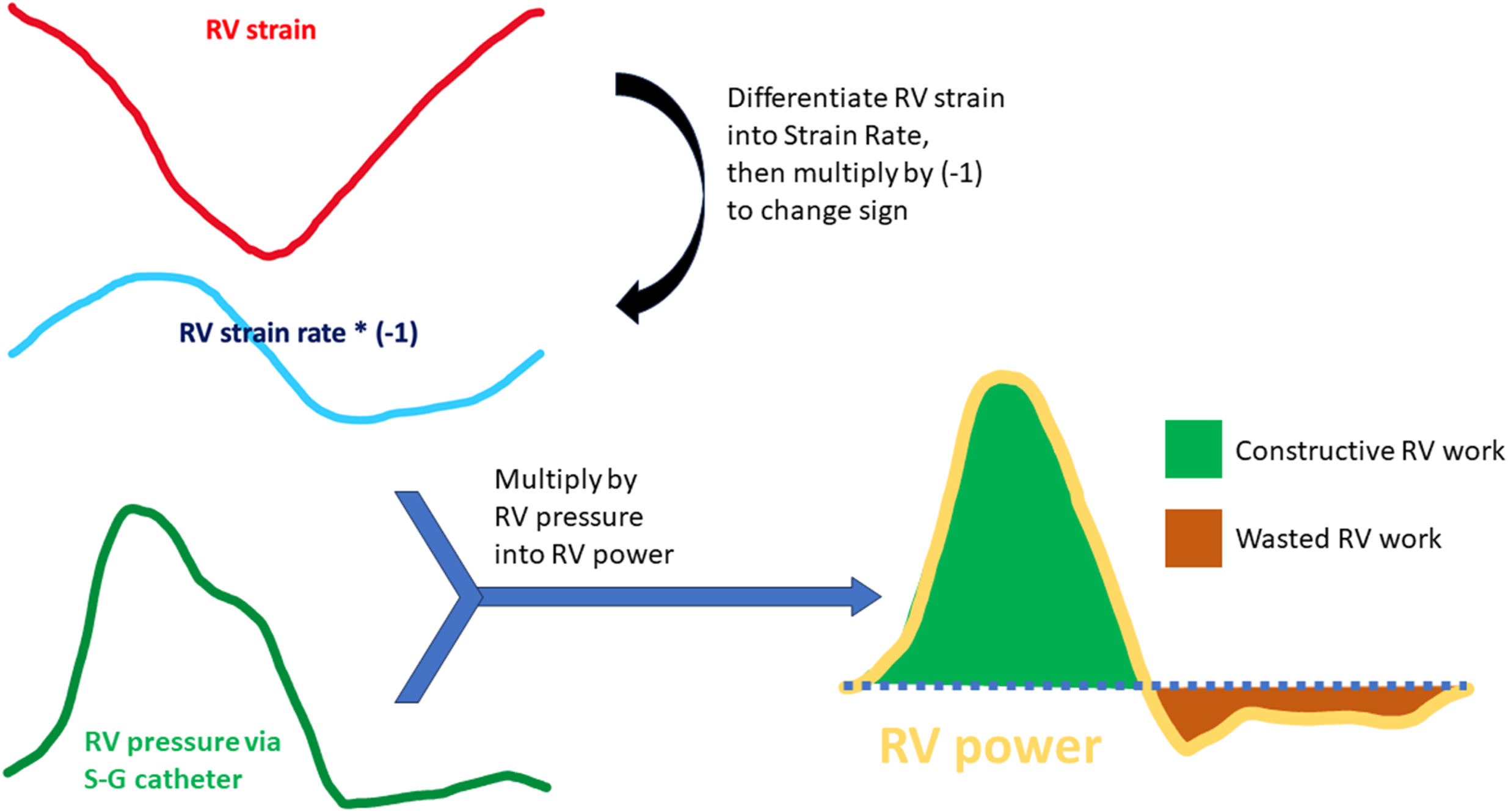
Method of Semi-Invasive Myocardial Work Analysis. Right ventricular (RV) strain curves (red) and RV strain rate curves (blue) are exported from dedicated software. RV pressure recordings obtained via a Swan-Ganz catheter, along with ECG signals, are averaged to create a single cardiac cycle RV pressure curve (from ECG R wave to R wave). Both the strain rate and RV pressure curves are resampled and then multiplied to generate the RV power curve. The positive area under the RV power curve represents constructive work (green), while the negative area represents wasted work (brown).

## RESULTS

Table 1 summarized the demographic characteristics, medication and basic laboratory data, and there was no significant difference between HFpEF and non-HFpEF groups.

**Table 1.** Patient characteristics.

|  | Non-HFpEF patients | HFpEF patients | p |
| --- | --- | --- | --- |
| n | 20 | 21 |  |
| age | 68.3±6.6 | 65.8±11.3 | 0.40 |
| Sex | 9/20 | 9/21 | 0.89 |
| BMI | 25.6±3.9 | 26.3±3.8 | 0.56 |
| BSA | 1.70±0.16 | 1.72±0.19 | 0.82 |
| H2FPEF score | 2.10 | 2.33 | 0.57 |
| Comorbidities |  |  |  |
| Coronary artery disease | 10 (50) | 11 (48) | 0.88 |
| Hypertension | 16 (80) | 13 (62) | 0.20 |
| Dyslipidemia | 10 (50) | 10 (48) | 0.88 |
| Diabetes mellitus | 5 (25) | 5 (23) | 0.92 |
| Medications |  |  |  |
| ACEI or ARB | 7 (35) | 5 (23) | 0.43 |
| Betablocker | 10 (50) | 13 (62) | 0.26 |
| CCB | 4 (20) | 3 (14) | 0.73 |
| statin | 8 (40) | 9 (43) | 0.65 |
| Diuretics | 4 (20) | 2 (10) | 0.41 |
| Nitrate | 4 (20) | 3 (14) | 0.73 |
| Laboratories |  |  |  |
| Hemoglobin | 13.7±0.9 | 13.5±1.5 | 0.63 |
| NT-proBNP | 109.1±133.2 | 389.6±1076.3 | 0.25 |
| HbA1bac | 6.0±1.1 | 5.9±0.6 | 0.84 |
| LDL | 100.9±32.6 | 103.5±31.3 | 0.80 |

ACEI, angiotensin-converting enzyme inhibitors; ARB, angiotensin receptor blockers;
BMI, body mass index; BSA, body surface area; CCB, calcium channel blocker;
HFpEF, heart failure with preserved ejection fraction; LDL, low density lipoprotein;
NT-proBNP, N-terminal pro- brain natriuretic peptide;

### Echocardiographic parameters (Table 2)

HFpEF groups had higher rest PCWP (21.2±6.4 vs. 12.9±3.4 mmHg, *p*<0.01), higher post-exercise PCWP (28.6±8.2 vs. 18.9±4.6 mmHg, *p*<0.01), higher mean PAP (28.1±7.4 vs. 20.5±4.9 mmHg, *p*<0.01) and higher post exercise men PAP (39.6±8.5 vs. 32.1±8.2 mmHg, *p*<0.01).

**Table 2.**
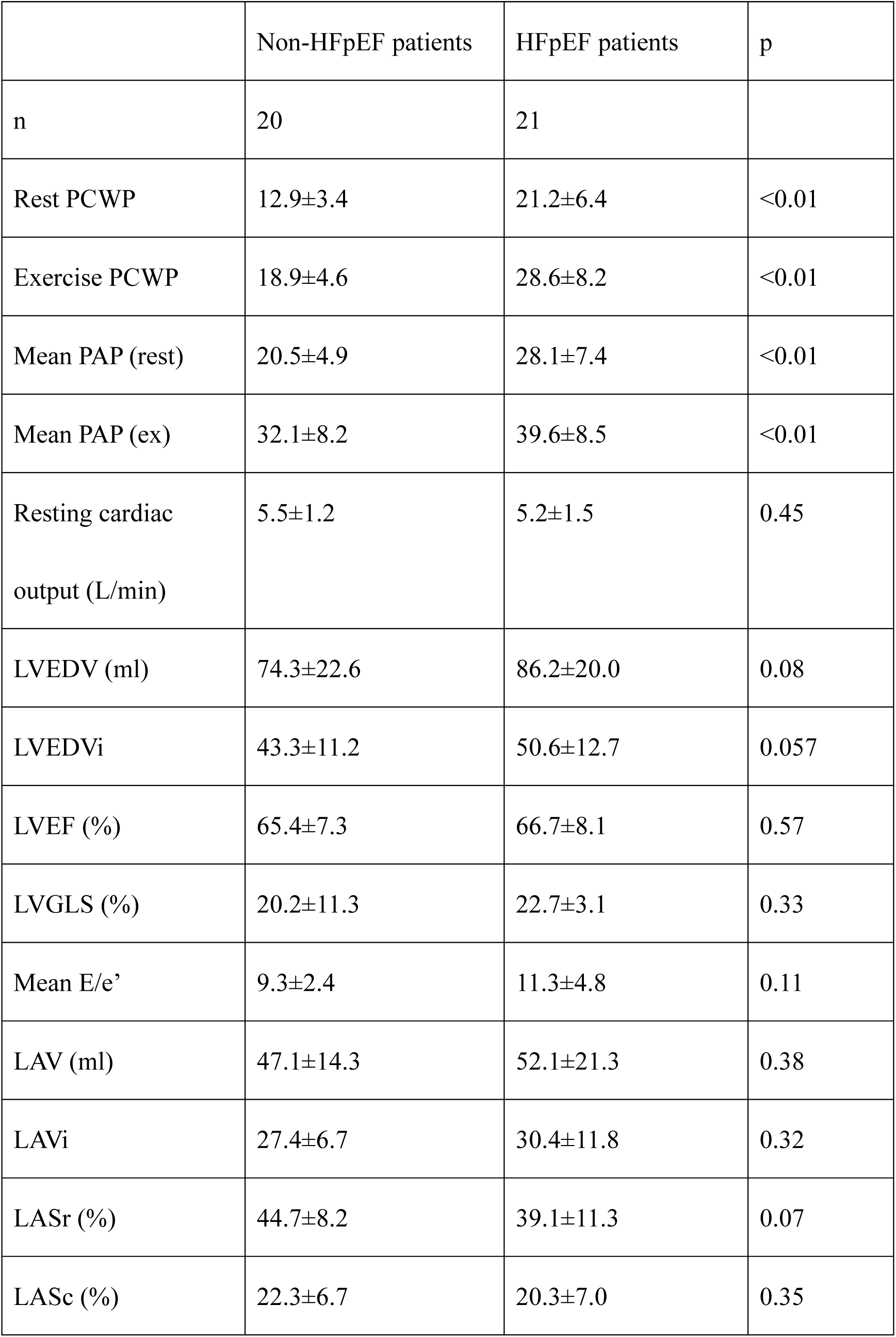
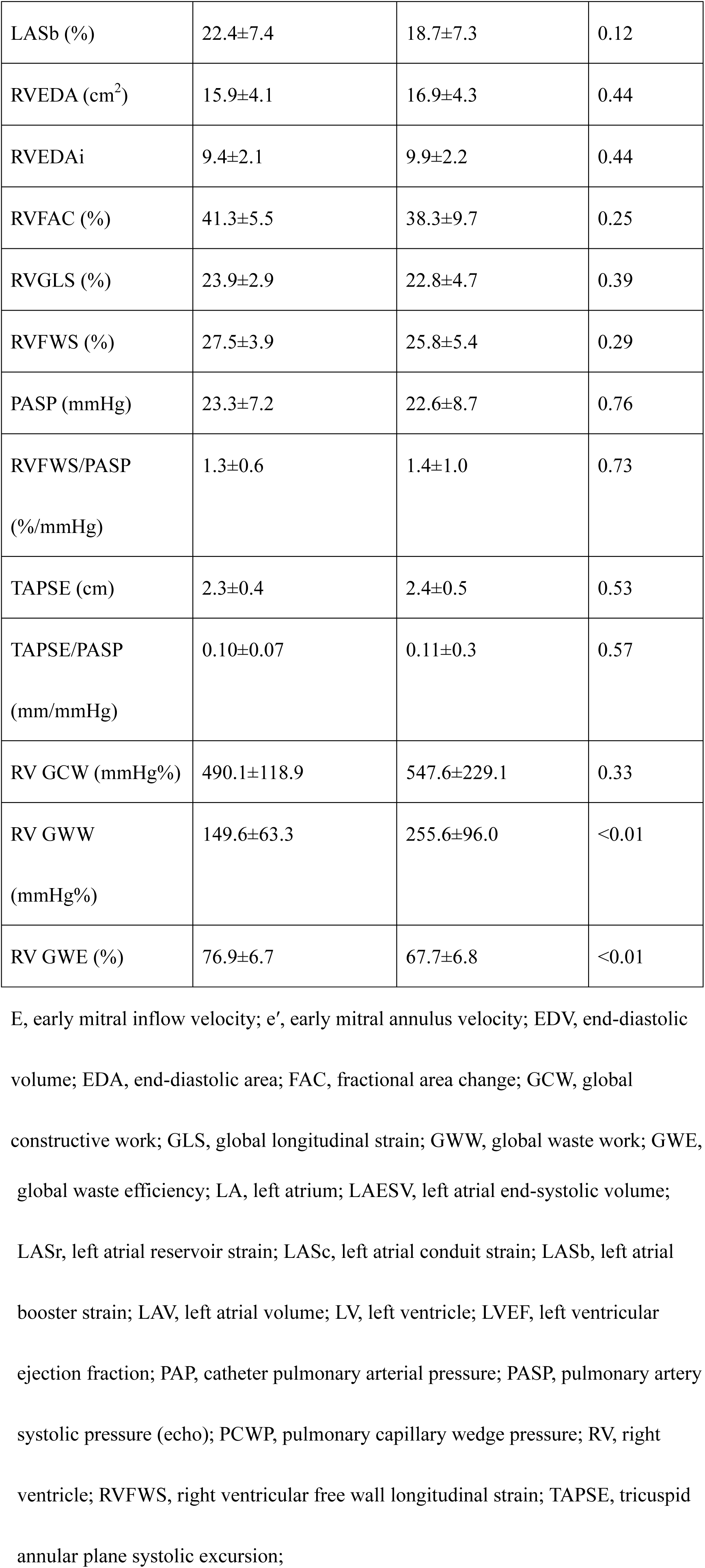
Echocardiographic and hemodynamic parameters.

|  | Non-HFpEF patients | HFpEF patients | p |
| --- | --- | --- | --- |
| n | 20 | 21 |  |
| Rest PCWP | 12.9±3.4 | 21.2±6.4 | <0.01 |
| Exercise PCWP | 18.9±4.6 | 28.6±8.2 | <0.01 |
| Mean PAP (rest) | 20.5±4.9 | 28.1±7.4 | <0.01 |
| Mean PAP (ex) | 32.1±8.2 | 39.6±8.5 | <0.01 |
| Resting cardiac output (L/min) | 5.5±1.2 | 5.2±1.5 | 0.45 |
| LVEDV (ml) | 74.3±22.6 | 86.2±20.0 | 0.08 |
| LVEDVi | 43.3±11.2 | 50.6±12.7 | 0.057 |
| LVEF (%) | 65.4±7.3 | 66.7±8.1 | 0.57 |
| LVGLS (%) | 20.2±11.3 | 22.7±3.1 | 0.33 |
| Mean E/e' | 9.3±2.4 | 11.3±4.8 | 0.11 |
| LAV (ml) | 47.1±14.3 | 52.1±21.3 | 0.38 |
| LAVi | 27.4±6.7 | 30.4±11.8 | 0.32 |
| LASr (%) | 44.7±8.2 | 39.1±11.3 | 0.07 |
| LASc (%) | 22.3±6.7 | 20.3±7.0 | 0.35 |
| LASb (%) | 22.4±7.4 | 18.7±7.3 | 0.12 |
| RVEDA (cm<sup>2</sup>) | 15.9±4.1 | 16.9±4.3 | 0.44 |
| RVEDA<sub>i</sub> | 9.4±2.1 | 9.9±2.2 | 0.44 |
| RVFAC (%) | 41.3±5.5 | 38.3±9.7 | 0.25 |
| RVGLS (%) | 23.9±2.9 | 22.8±4.7 | 0.39 |
| RVFWS (%) | 27.5±3.9 | 25.8±5.4 | 0.29 |
| PASP (mmHg) | 23.3±7.2 | 22.6±8.7 | 0.76 |
| RVFWS/PASP<br>(%/mmHg) | 1.3±0.6 | 1.4±1.0 | 0.73 |
| TAPSE (cm) | 2.3±0.4 | 2.4±0.5 | 0.53 |
| TAPSE/PASP<br>(mm/mmHg) | 0.10±0.07 | 0.11±0.3 | 0.57 |
| RV GCW (mmHg%) | 490.1±118.9 | 547.6±229.1 | 0.33 |
| RV GWW<br>(mmHg%) | 149.6±63.3 | 255.6±96.0 | <0.01 |
| RV GWE (%) | 76.9±6.7 | 67.7±6.8 | <0.01 |

E, early mitral inflow velocity; e', early mitral annulus velocity; EDV, end-diastolic volume; EDA, end-diastolic area; FAC, fractional area change; GCW, global constructive work; GLS, global longitudinal strain; GWW, global waste work; GWE,
global waste efficiency; LA, left atrium; LAESV, left atrial end-systolic volume; LASr, left atrial reservoir strain; LAsC, left atrial conduit strain; LASb, left atrial booster strain; LAV, left atrial volume; LV, left ventricle; LVEF, left ventricular ejection fraction; PAP, catheter pulmonary arterial pressure; PASP, pulmonary artery systolic pressure (echo); PCWP, pulmonary capillary wedge pressure; RV, right ventricle; RVFWS, right ventricular free wall longitudinal strain; TAPSE, tricuspid annular plane systolic excursion;

For left heart echocardiographic parameters, there was no significant difference in LVEDV (86.2±20.0 vs. 74.3±22.6 ml, *p*=0.08), LVEF (66.7±8.1 vs. 65.4±7.3 %, *p*=0.57), LVGLS (22.7±3.1 vs. 20.2±11.3, *p*=0.33), mean E/e’ (11.3±4.8 vs. 9.3±2.4, *p*=0.11), LAV (52.1±21.3 vs. 47.1±14.3 ml, *p*=0.38), LASr (39.1±11.3 vs. 44.7±8.2%, *p*=0.07), LASc (20.3±7.0 vs. 22.3±6.7%, *p*=0.35), and LASb (18.7±7.3 vs. 22.4±7.4%, *p*=0.12).

For right heart echocardiographic parameters, there was no significant difference in RVEDA (16.9±4.3 vs. 15.9±4.1 cm^2^, *p*=0.44), RVFAC (38.3±9.7 vs. 41.3±5.5 %, *p*=0.25), RVGLS (22.8±4.7 vs. 23.9±2.9 %, *p*=0.39), RVFWS (25.8±5.4 vs. 27.5±3.9 %, *p*=0.29), TAPSE (2.4±0.5 vs. 2.3±0.4 cm, *p*=0.53), RVFWS/PASP (1.4±1.0 vs. 1.3±0.6 %/mmHg, *p*=0.73), and TAPSE/PASP (1.0±0.3 vs. 1.2±0.7 cm/mmHg, *p*=0.57).

### Right ventricular myocardial work analysis (Table 2 and Figure 2)

For right ventricular myocardial work analysis, there was no significant difference in RVGCW (547.6±229.1 vs. 490.1±118.9 mmHg%, *p*=0.33). But HFpEF groups had higher RV GWW (255.6±96.0 vs. 149.6±63.3 mmHg%, *p*<0.01) and lower RV GWE (67.7±6.8 vs. 76.9±6.7%, *p*<0.01).

**Figure 2.**
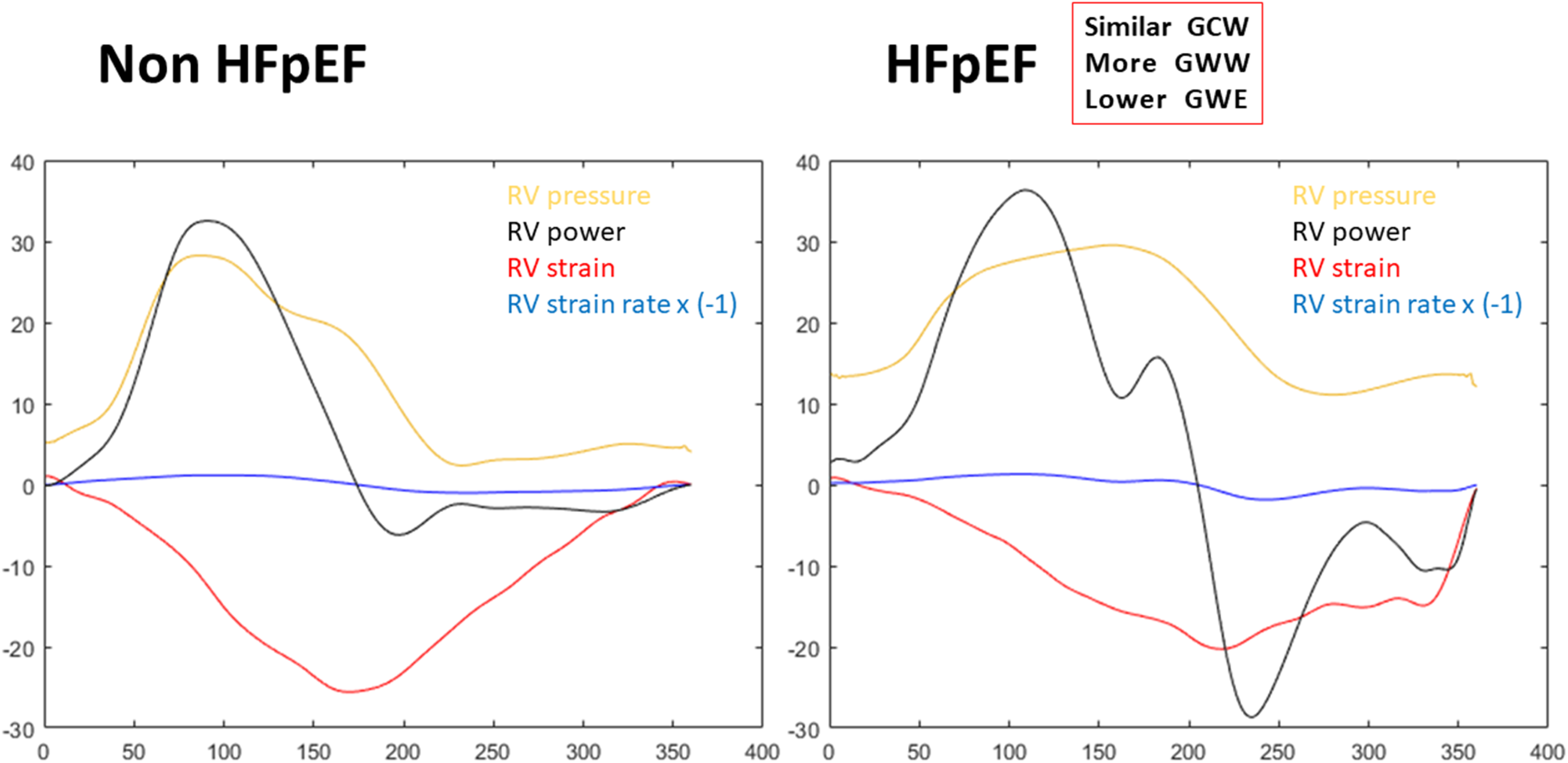
Right Ventricular Myocardial Work Analysis. Curves of RV pressure (yellow), RV strain (red), inversed RV strain rate (blue) and RV power (black) are plotted together for a heart failure with preserved ejection fraction (HFpEF) patient (right) and a non-HFpEF patient (left). Peak pressure and peak RV shortening are delayed and the RV wasted work (the negative area of the RV power curve) is more in the HFpEF patient.

Figure 3 reveals that RV GWW is significantly correlated with rest PCWP (r=0.48, *p* <0.01), post-exercise ΔPCWP (r=0.41, *p* <0.01), but not LVSVi (r=-0.24, p=0.13) or cardiac index (r=-0.05, p=0.76).

**Figure 3.**
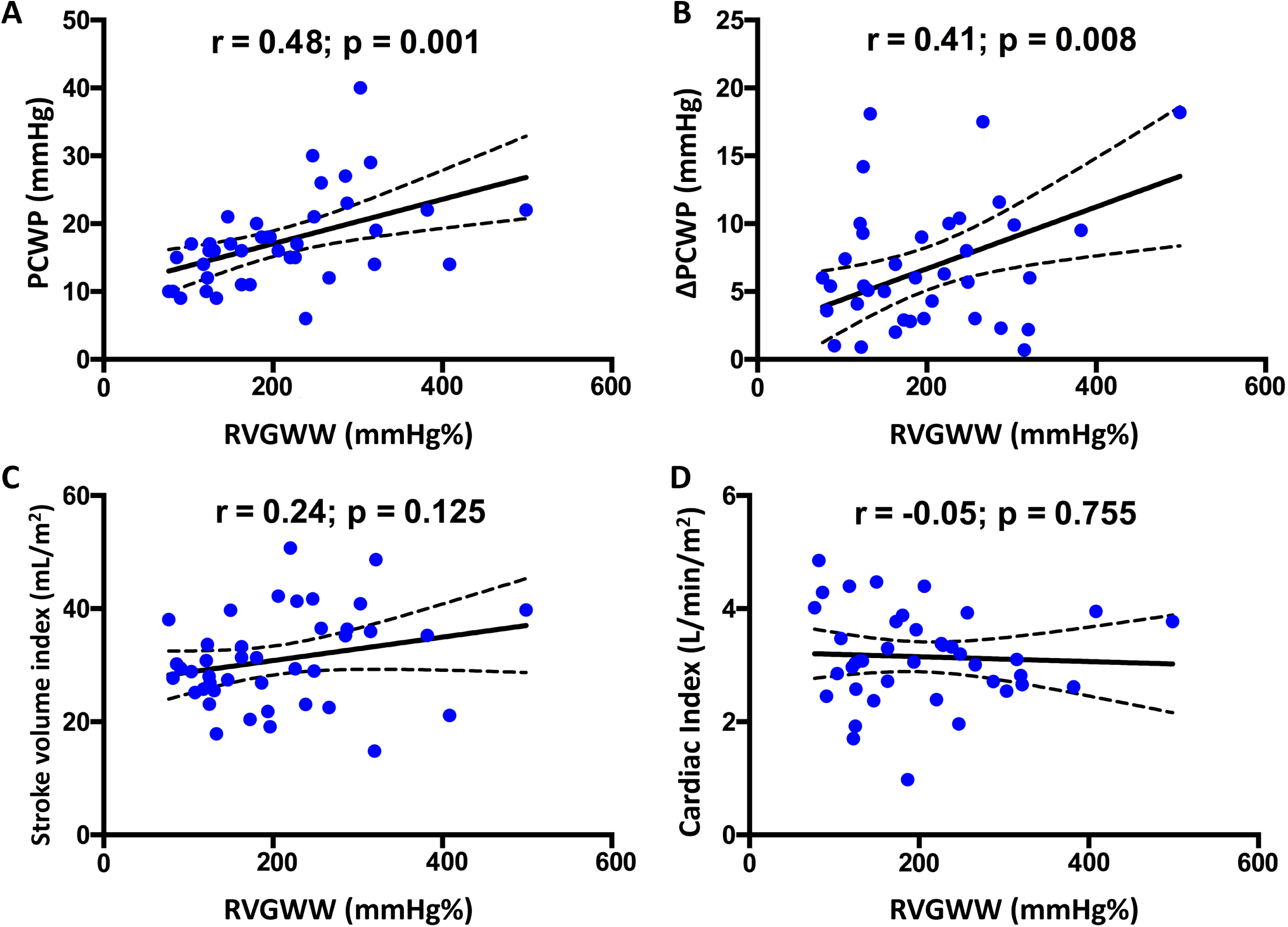
Correlation of RVGWW with Parameters from Invasive. **Cardiopulmonary Exercise Test** Significant correlations between RVGWW and invasively derived rest PCWP, as well as post-exercise ΔPCWP, are evident. However, no significant correlations are observed with stroke volume index and cardiac index. **Central illustration** Right Ventricular Myocardial Work in Heart Failure with Preserved Ejection Fraction. Right ventricle global waste work (RVGWW) is significantly increased in HFpEF patients and correlated with the change of PCWP at peak exercise. Abbreviations : HFpEF, heart failure with preserved ejection fraction; iCPET, invasive cardiopulmonary exercise test; RV, right ventricle; PCWP, pulmonary capillary wedge pressure.

### Prediction of HFpEF with echocardiographic parameters (Table 3)

To predict the diagnosis of HFpEF via iCPET, the RV GWW and RV GWE had higher AUC (0.84 and 0.83, respectively) than mean E/e’ (AUC 0.59), LASr (AUC 0.64), RVFAC (AUC 0.61), TAPSE/PASP (AUC 0.50) and RVFWS/PASP (AUC 0.52).

**Table 3.** Prediction of HFpEF with echocardiographic parameters.

| HFpEF | AUC | 95% C.I. | Sensitivity (%) | Specificity (%) | Diagnostic accuracy (%) | Cut-off |
| --- | --- | --- | --- | --- | --- | --- |
| RVGWW | 0.85 | 0.73-0.97 | 95 | 57 | 92 | 173 |
| RVGWE | 0.83 | 0.70-0.96 | 80 | 86 | 84 | 74 |
| RVGCW | 0.59 | 0.41-0.77 | 50 | 76 | 67 | 453 |
| Averaged E/e' | 0.59 | 0.41-0.77 | 55 | 42 | 43 | 10.3 |
| E/A | 0.5 | 0.31-0.69 | 60 | 40 | 52 | 0.88 |
| LASr | 0.64 | 0.47-0.81 | 90 | 38.1 | 58 | 35 |
| LAVi | 0.57 | 0.39-0.75 | 70 | 24 | 55 | 31.5 |
| RVFAC | 0.61 | 0.43-0.79 | 80 | 52 | 54 | 37.2 |
| RVFWS | 0.60 | 0.42-0.79 | 85 | 52 | 50 | 24.6 |
| TAPSE/PASP | 0.50 | 0.30-0.70 | 88.2 | 40 | 62 | 0.23 |
| RVFWS/PASP | 0.52 | 0.33-0.7 | 40 | 75 | 61 | 1.35 |

Abbreviations as in Table 2

We also examined the relationship of echocardiographic parameters and RV myocardial work with post-exercise ΔPCWP and exercise PCWP (Supplemental Table 1). LASb was negatively correlated and RV GCW and GWW were positively correlated with post-exercise ΔPCWP. LASr, LASb and RV GWE were negatively correlated with exercise PCWP. And mean E/e’, RV GCW and GWW were positively correlated with exercise PCWP.

We performed multivariate analysis to control for possible confounding factors (Table 4), including age, LVGLS, LAVi, LASc and mean E/e’. The β coefficient for every 100 mmHg% of RV GWW was 1.981 (0.056∼3.906, *p*=0.044) for post-exercise ΔPCWP and 4.323 (2.429∼6.218, *p*<0.001) for exercise PCWP after adjustment.

**Table 4.** Multivariate regression analyses with post-exercise ΔPCWP and exercise PCWP as the dependent variables.

| | Post-exercise ΔPCWP | | | exercise PCWP | | |
| --- | --- | --- | --- | --- | --- | --- |
| | $\beta$ (95% CI) | | p value | $\beta$ (95% CI) | | p value |
| RV<br>GWW* | 1.981 | 0.056~3.906 | 0.044 | 4.323 | 2.429~6.218 | <0.001 |
| age | 0.033 | -0.186~0.253 | 0.759 | 0.036 | -0.178~0.250 | 0.734 |
| LVGLS | -0.007 | -0.216~0.202 | 0.945 | 0.122 | 0.081~0.325 | 0.229 |
| LAVi | 0.026 | -0.180~0.232 | 0.799 | 0.130 | -0.078~0.337 | 0.209 |
| LASc | 0.284 | -0.163~0.365 | 0.443 | -0.145 | -0.115~0.405 | 0.263 |
| Mean E/e' | -0.101 | -0.184~0.752 | 0.226 | 0.918 | 0.463~1.373 | <0.001 |

Abbreviations as in Table 2; \* $\beta$ value per 100 mmHg

## DISCUSSION

Exercise intolerance and dyspnea in patients with HFpEF can be attributed to exercise-induced dynamic congestion caused by rapidly increased PCWP. The elevated afterload and RV dysfunction sustain challenges to cardiac dynamics through an unfavorable RV to LV diastolic interaction ^20^. The present study suggests that pressure-strain loop analysis of RVMW improves the accuracy of HFpEF diagnosis. Compared with echocardiographic parameters such as averaged E/e’, LAVI, and strain, RV GWW and RV GWE demonstrate better predictive ability for diagnosing HFpEF. Additionally, increased RV GWW is significantly correlated with PCWP and ΔPCWP during exercise. Pressure-strain loop analysis of our HFpEF cohort provides insights into understanding the pathophysiology of patients with HFpEF.

### Application of myocardial work in RV pathophysiology

The right ventricle is no longer a forgotten chamber. Whenever a new assessment method for the LV emerges, it is quickly extrapolated for RV application. Techniques such as 3D volumetric RVEF, speckle tracking deformational analysis for RV longitudinal strain, and ventricular arterial coupling concepts now have RV-specific versions (i.e. TAPSE/PASP, RVFWS/PASP). These traditional parameters typically represent changes in the heart from end-systole to end-diastole, which are useful for the LV, a pumping chamber, but not entirely suitable for the RV, a peristalsis conduit chamber. The pulmonary artery Doppler w-shape in pulmonary hypertension and the delay in RV peak pressure or contraction ^21,22^ both underscore the need to consider temporal effects and such unique mechanics in RV function evaluation.

Pressure-strain analysis is a remarkable invention. Unlike pressure-volume analysis, by differentiating strain into strain rate, more inflection points are introduced in the curve (Figure 1). This allows the integration of pressure to derive the concept of wasted work. Following Butcher’s pioneering application of myocardial work analysis to the RV, many teams have validated non-invasive RVMW in various diseases ^23–25^. Our team was the first to apply RVMW in a HFpEF population confirmed with iCPET, and found that RV GWW effectively explained and predicted changes in exercise PCWP.

Due to the difficulty in obtaining ideal tricuspid regurgitation doppler envelope in our patient population, we adopted a semi-invasive rather than a fully non-invasive approach to perform RVMW analysis. Future improvements in non-invasive RVMW analysis could come from establishing RV pressure reference curves and using ultrasound contrast agents to optimize RV free wall tracking ability and doppler signal quality.

### Superiority of RVMW over other parameters of RV and pulmonary circulation (Pc) uncoupling

PCWP is an essential hemodynamic parameter in HFpEF, reflecting left ventricular diastolic function and influencing symptoms and prognosis in affected patients. Elevated PCWP can increase RV afterload, making the RV stiffer and impairing its filling and contractile properties, leading to uncoupling with pulmonary circulation.^26^ Pressure-strain loops theoretically provide a more comprehensive evaluation of RV function through the calculation of RVMW indices, compared to standard echocardiographic measures.^11^ Unlike RV longitudinal strain, TAPSE, and RV FAC, RVMW parameters incorporate contractility, RV dyssynchrony, and pulmonary pressures into their assessment. ^15^ This comprehensive evaluation of RV function is not affected by the technical limitations present in other standard parameters. TAPSE’s measurement is influenced by angle and load and varies with cardiac translation, while RV FAC’s reliability is hindered by increased load dependency and only modest interobserver reproducibility.^27^ RV longitudinal strain is affected by afterload. By accounting for afterload in the assessment, RVMW provides insight into RV-pulmonary arterial coupling, potentially offering a more accurate evaluation of PCWP and LVEDP.^15^ RVMW integrates RV dyssynchrony and post-systolic shortening into the evaluation of RV function by synchronizing pulmonic and tricuspid valvular events with RV longitudinal strain. For patients with HFpEF, both cardiac and extracardiac components contribute to limited exercise capacity, which can lead to RV and Pc uncoupling, dyssynchronous left-to-right delay, and ventricular interdependence. ^11,20,22,28^ In patients with HFrEF, RVGCW demonstrated a moderate correlation with invasively measured stroke volume and stroke volume index.^15^ Additionally, RV global wasted work (RV GWW) represents myocardial lengthening during systole and shortening during isovolumic relaxation, which do not contribute to RV constructive work. Our study showed that increased RV GWW improves the diagnostic accuracy of HFpEF and moderately correlates with PCWP, though it is not related to stroke volume and cardiac index.

### Clinical implication

Among myocardial work metrics, LVGCW was found to be a superior indicator of exercise capacity compared to global longitudinal strain in a cohort of 114 patients with HFpEF. The increase in LVGCW observed during exercise in patients treated with spironolactone for six months is believed to correlate with improvements in functional capacity. ^29^ RVGCW, derived from non-invasive assessment, emerges as a superior parameter, offering an integrated analysis of RV systolic function. It correlates more strongly with invasively measured stroke volume and stroke volume index compared to other standard echocardiographic parameters.^15^ In this study, we have demonstrated that RVMW parameters provide a comprehensive assessment of PCWP and ΔPCWP during exercise in individuals with HFpEF. By combining RV strain with invasive right heart catheterization for strain-pressure loop analysis, we can effectively characterize various RV pathologies and regional RV myocardial energetics in HFpEF. Our findings highlight the impact of increased LVEDP and PCWP on RV dyssynchrony, ventricular interdependence, and wasted RV work. Additionally, the novel parameter RVGWW could predict the rise in PCWP during exercise and explain the exercise intolerance observed in HFpEF patients. Our approach suggests that the need for exercise testing may be eliminated while maintaining the accuracy of HFpEF diagnosis.

### Limitations

The major limitation of this proof-of-concept study is that the pressure-strain analysis utilized is not based on current commercial software. Consequently, our approach is not entirely non-invasive, as RV pressure recordings still depend on measurements obtained from a Swan-Ganz catheter. Currently available commercial software enables the non-invasive assessment of LV myocardial work from the estimated LV pressure-time curve via adjusting the empiric reference LV pressure curve. The crucial step is to use measured systolic and diastolic blood pressures from a sphygmomanometer and to use echocardiography derived isovolumic contraction time (IVCT), ejection time, and isovolumic relaxation time (IVRT) for the simulation of an individualized LV pressure-time curves.

For the RV, such dedicated empiric reference ventricular pressure curve is lacking ^30^. On the other hand, as the typical RV pressure-volume curve is triangular but not rectangular ^31^, it is hard to precisely define the IVCT and IVRT of RV. In conditions such as pulmonary hypertension or severe left heart congestion, the RV pressure-volume curve can somewhat like their left counterpart, and the adoption of the commercial LV myocardial work analysis software might be feasible. Butcher and colleagues have successfully demonstrated that the RV GWW were significantly higher in HFrEF and pulmonary hypertension patients via such non-invasive pressure-strain analysis ^15,32^. However, it is difficult to perform a satisfactory non-invasive RV pressure estimation in our HFpEF patients. Instead, we used ECG signals as a reference to conjugate the invasive pressure curve and the non-invasive strain curves for pressure-strain analysis. The applicability of our approach to other pathophysiological conditions, such as HFrEF, mitral valve disease, aortic valve disease, or pulmonary arterial hypertension, remains to be validated in subsequent studies.

## CONCLUSION

We developed a vendor-independent semi-invasive myocardial work analysis method by combining RV strain with RV pressure recordings and validated it using iCPET-confirmed HFpEF. Our findings indicate that RV GWW not only effectively predicts HFpEF but also better predicts exercise PCWP and post-exercise ΔPCWP compared to other standard echocardiographic parameters in HFpEF.

## Nonstandard Abbreviations and Acronyms

ΔPCWP: Change in Pulmonary Capillary Wedge Pressure
GCW: Global Constructive Work
GWE: Global Work Efficiency
GWI: Global Work Index
GWW: Global Wasted Work
HFpEF: Heart Failure with Preserved Ejection Fraction
HFrEF: Heart Failure with Reduced Ejection Fraction
iCPET: Invasive Cardiopulmonary Exercise Test
IVCT: Isovolumic Contraction Time
IVRT: Isovolumic Relaxation Time
LAV: Left Atrial Volume
LAVI: Left Atrial Volume Index
LASb: Left Atrial Booster Strain
LASc: Left Atrial Conduit Strain
LASr: Left Atrial Reservoir Strain
LV: Left Ventricle / Left Ventricular
LVEDP: Left Ventricular End-Diastolic Pressure
LVEDV: Left Ventricular End-Diastolic Volume
LVESV: Left Ventricular End-Systolic Volume
LVGLS: Left Ventricular Global Longitudinal Strain
LVEF: Left Ventricular Ejection Fraction
LVSV: Left Ventricular Stroke Volume
NT-proBNP: N-terminal pro B-type Natriuretic Peptide
PA: Pulmonary Artery
PAP: Pulmonary Artery Pressure
PCWP: Pulmonary Capillary Wedge Pressure
RA: Right Atrium / Right Atrial
RV: Right Ventricle / Right Ventricular
RVEDA: Right Ventricular End-Diastolic Area
>VESA: Right Ventricular End-Systolic Area
RVFAC: Right Ventricular Fractional Area Change
RVFWS: Right Ventricular Free Wall Strain
RVGCW: Right Ventricular Global Constructive Work
RVGLS: Right Ventricular Global Longitudinal Strain
RVGWE: Right Ventricular Global Work Efficiency
RVGWI: Right Ventricular Global Work Index
RVGWW: Right Ventricular Global Wasted Work
RVMW: Right Ventricular Myocardial Work
SV: Stroke Volume
SVi: Stroke Volume Index
TAPSE: Tricuspid Annular Plane Systolic Excursion

## Declarations

### Data Availability Statement

No new data were generated or analyzed in support of this research. The datasets used in this study were only available in the National Taiwan University Hospital.

### Ethics approval and consent to participate

The study protocol complies with the Declaration of Helsinki and was approved by the Institutional Review Board of National Taiwan University Hospital (IRB 201908057RINC).

### Consent for publication

Not applicable.

### Authors’ contributions

TT-L and LY-L contributed to the conception or design of the work and analyzed data. LY-L, CK-W, and KC-H contributed to the acquisition of data for the work. KC-H and TT-L drafted and revised the manuscript. TT-L and CK-W critically revised the manuscript. All gave final approval and agreed to be accountable for all aspects of work ensuring integrity and accuracy.

---

**Supplemental Figure 1.**
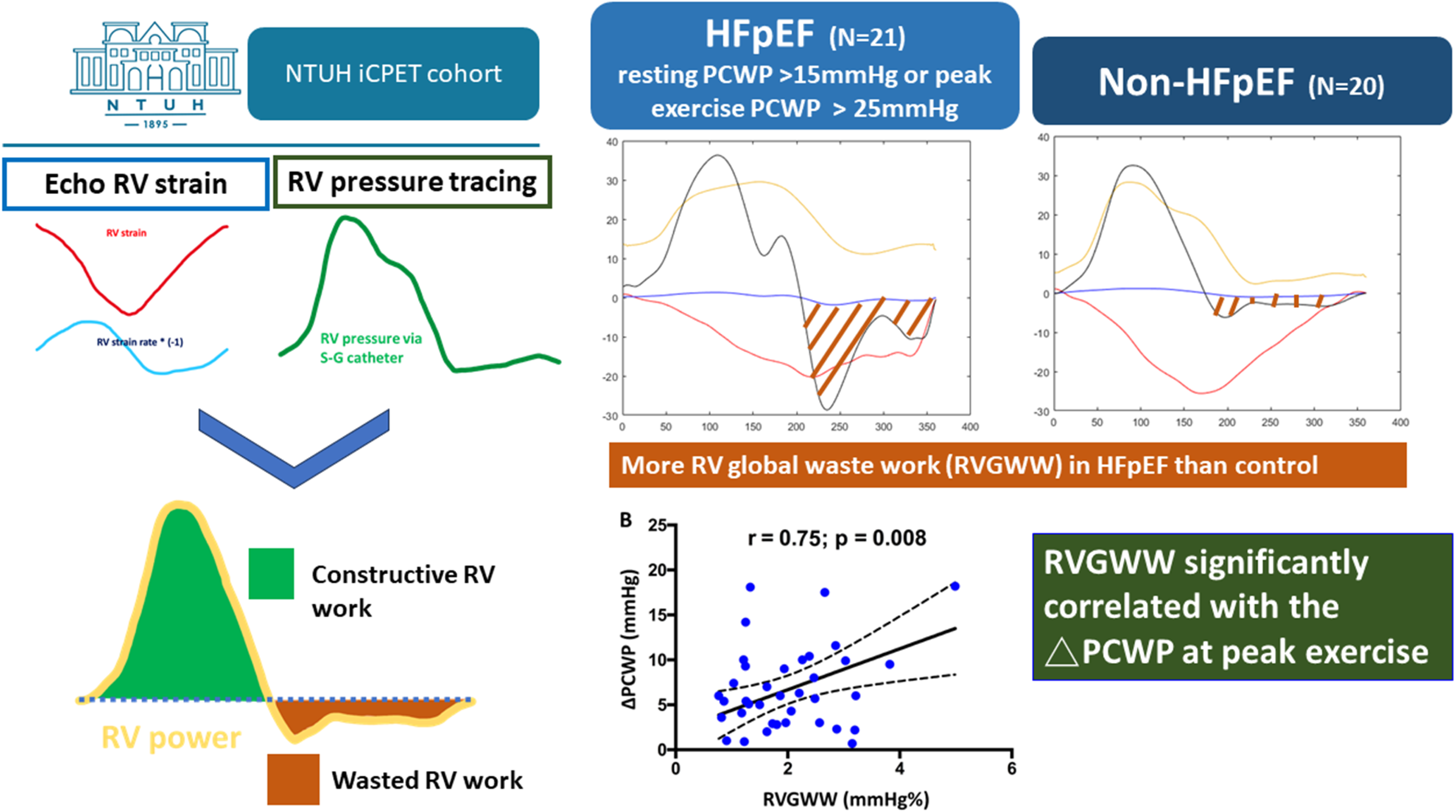
The Patient Enrollment Algorithm.

